# Proteome-wide autoantibody screening and holistic autoantigenomic analysis unveil COVID-19 signature of autoantibody landscape

**DOI:** 10.1101/2024.06.07.24308592

**Authors:** Kazuki M Matsuda, Yoshiaki Kawase, Kazuhiro Iwadoh, Makoto Kurano, Yutaka Yatomi, Koh Okamoto, Kyoji Moriya, Hirohito Kotani, Teruyoshi Hisamoto, Ai Kuzumi, Takemichi Fukasawa, Asako Yoshizaki-Ogawa, Masanori Kono, Tomohisa Okamura, Hirofumi Shoda, Keishi Fujio, Kei Yamaguchi, Taishi Okumura, Chihiro Ono, Yuki Kobayashi, Ayaka Sato, Ayako Miya, Naoki Goshima, Rikako Uchino, Yumi Murakami, Hiroshi Matsunaka, Hiroshi Imai, Shinichi Sato, Rudy Raymond, Ayumi Yoshizaki

## Abstract

This study presents “aUToAntiBody Comprehensive Database (UT-ABCD)”, a comprehensive catalog of autoantibody profiles in 284 human individuals. The subjects include patients diagnosed with Coronavirus disease 2019 (COVID-19; n = 73), systemic sclerosis (SSc; n = 32), systemic lupus erythematosus (SLE; n = 60), anti-neutrophil cytoplasmic antibody-associated vasculitis (AAV; n = 29), atopic dermatitis (AD; n = 26), as well as healthy controls (HC; n = 64). Our investigation employs proteome-wide autoantibody screening (PWAbS) that utilizes 13,352 autoantigens displayed on wet protein arrays (WPAs). Our WPAs display human proteins synthesized *in vitro* utilizing a wheat germ cell-free system, maintained in a hydrated state. Our findings demonstrated significant elevation in the number of IgG autoantibody positivity in COVID-19, SSc, SLE, AAV, and AD patients compared to HCs. Employing machine learning, we distinguished COVID-19 cases with high accuracy based on autoantibody profiles, notably identifying antibodies against proteins encoded by *BCORP1* and *KAT2A* as highly specific to COVID-19 (specificity: 87% and 97%, respectively). Our research highlights the effectiveness of integrating PWAbS and autoantigenomics in exploring immune responses in COVID-19 and other diseases. It provides a deeper understanding of the autoimmunity landscape in human disorders and introduces a new bioresource for further investigation.

## Introduction

Coronavirus disease 2019 (COVID-19), an infectious disease caused by severe acute respiratory syndrome coronavirus 2 (SARS-CoV-2),^1^ has brought a global pandemic since early 2020 with threat on human health and public safety throughout the world.^2^ The pathophysiology of COVID-19 is characterized by multiple organ injuries triggered by excessive immune response.^3,4^ Cytokine storm in the lung causes acute respiratory distress syndrome, which leads to hypoxemia, respiratory failure, requirement of ventilation, and even death. One of the biggest challenges in clinical management of COVID-19 patients lies in accurately identifying and categorizing cases at higher risk of such serious clinical course. Known risk factors include older age, male gender, smoking, diabetes, obesity, hypertension, immunodeficiency, and malignancies.^5^

Humoral immunity plays pivotal roles in COVID-19. Although dramatic success of mRNA vaccines and SARS-CoV-2 neutralizing monoclonal antibodies in preventing serious illnesses, accumulating evidence have suggested the vicious roles of dysregulated humoral immunity.^6–10^ As well as earlier work linking anti-cytokine antibodies to mycobacterial, staphylococcal and fungal diseases,^11,12^ autoantibodies against cytokines have been described in COVID-19.^13^ Especially, anti-type I Interferon antibodies distinguished ∼10% of life-threatening pneumonia and ∼20% of deaths from COVID-19.^6–8^ A high-throughput screening by yeast display of the secretome further revealed the presence of autoantibodies against several immune factors, including chemokines.^14^ In addition, autoantibodies characteristic of systemic autoimmune disorders, such as anti-phospholipid antibodies, anti-nuclear antibodies and rheumatoid factor, were reported in COVID-19.^15^ More recently, association between COVID-19 severity and autoantibodies targeting G protein-coupled receptors and renin-angiotensin system-related proteins has been reported.^16^

To comprehensively understand such clinical significance of autoantibodies in human diseases including COVID-19, high-precision autoantibody measurement with a proteome-wide scale is necessary. Herein, we employed an original protein microarray technology, which includes over 13,350 proteins, for proteome-wide autoantibody screening (PWAbS),^17–19^ in the serum samples derived from individual patients. Our pipeline integrates human cDNA library (HuPEX),^20^ a wheat germ cell-free system for high-throughput *in vitro* protein synthesis,^21–23^ and technology for manipulating protein arrays kept in moist conditions during the entire handling process,^24^ namely wet protein arrays (WPAs). We have applied this method in multiple inflammatory or malignant disorders for validating its potential for illustrating the “autoantibody landscape” of human disorders, which revealed its usefulness for holistic evaluation of disease-related autoantibodies,^19^ developing novel biomarkers,^17^ and moreover, investigating unknown pathophysiology driven by autoantibodies.^18^ While the significant body of work that has utilized proteome-wide approaches to investigate autoantibodies in the context of COVID-19, ^9,10^ our study provides a complementary perspective by a focus on a Japanese cohort, use of a different technology, comparison with a variety of conditions other than COVID-19, and longitudinal analysis.

Our aim was to demonstrate the utility of our omics-based methodology for autoantibody evaluation and data interpretation procedure, so-called “autoantigenomics,” targeting COVID-19. In 2020, Moritz *et al.* defined autoantigenomics as a branch of systems immunology, which holistically analyze the repertoire of autoantibodies engaging omics-based bioinformatical approaches including hierachical cluster analysis, enrichment analysis, and machine learning.^25^ The concept of autoantigenomics stand on hypotheses that there might be differences in the sets of targeted antigens underlying intra-disease heterogeneity in human, which would be supported by our novel data shown below.

## Results

### Overview

We recruited 73 patients with COVID-19, 32 patients with systemic sclerosis (SSc), 60 patients with systemic lupus erythematosus (SLE), 29 patients with anti-neutrophil cytoplasmic antibody-associated vasculitis (AAV), 26 patients with atopic dermatitis (AD), and 64 healthy controls (HC) for serum sample collection (**Extended Table 1**). For each individual serum, PWAbS was performed focusing on IgG autoantibodies (**Fig. 1A**). Our WPA included 84.8% of previously reported housekeeping genes (**Fig. 1B**).^26^ Based on genes expressed at more than 3.5 transcripts per million (TPM) in the Genotype-Tissue Expression (GTEx) V8,^27^ our WPA covered 72.1% of expressed human genes (**Fig. 1C**).

**Figure 1.**
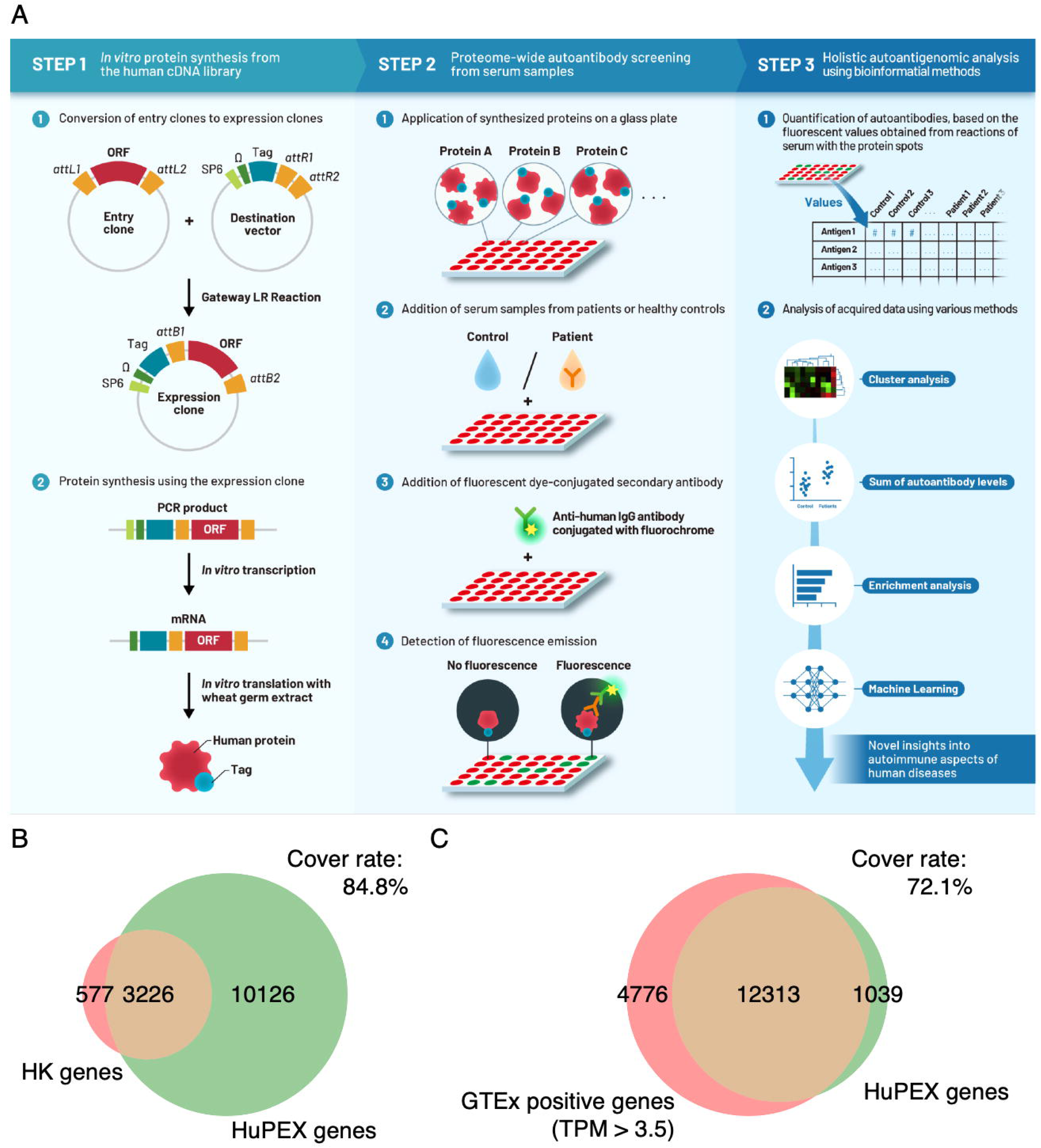
Overview of PWAbS methodology. **(A)** Scheme of PWAbS pipeline. In the first step, proteins were synthesized *in vitro* from the proteome-wide human cDNA library (HuPEX). Promotors (P), Enhancers (E), and FLAG-GST tags were fused to open reading frames of the expression clones by Gateway LR reaction. After polymerase chain reaction amplification and *in vitro* transcription, translation was performed using the wheat germ cell-free synthesis system. In the second step, we prepared WPAs by plotting synthesized proteins onto glass slides in an array format. WPAs were treated with serum samples derived from diseased patients or HCs. Autoantibodies were detected by fluorochrome-conjugated anti-human IgG Ab. In the third step, autoantibody quantification was performed based on the fluorescent values. Analysis of acquired high-dimensional autoantibody profiles was conducted by multiple omics-based approaches. ORF: open reading frame. (B) A venn diagram shows our WPA covers 84.8% of human housekeeping genes. (C) A venn diagram illustrates our WPA covers 72.1% of expressed human genes, based on genes expressed at more than 3.5 TPM in the GTEx V8, a bulk RNA-seq data from 52 human tissues and two cell lines.

The digest of the results is available as “aUToAntiBody Comprehensive Database (UT-ABCD)” at http://www.ut-abcd.org. We found that sum of autoantibody levels (SAL), defined as the sum of all quantified values for the 13,352 displayed autoantigens, was significantly elevated in patients with COVID-19 (median [interquartile range]: 4418 [3325-5392] AU), SSc (5784 [3960-7338] AU), or SLE (5963 [4543-7386] AU), compared to HCs (3014 [2289-3851] AU), while there was no statistically significant difference in SAL between AD (3664 [2419-5451] AU) or AAV (3842 [3281-4547] AU) patients and HCs (**Extended Fig. 2A**). This tendency was consistent across both gender and age groups. (**Extended Fig. 2B**).

### Threshold Determination and Positivity Counts

To establish the threshold for determining serum positivity of each autoantibody, Z-scores were calculated based on the mean and standard deviation values among healthy controls (HCs). Analysis revealed that over 99% of Z-scores in HCs were below 4 (**Extended Fig. 1C, 1D, and 1E**). Consequently, a threshold of Z-scores > 4 was set to define autoantibody seropositivity. The number of positive autoantibodies was then counted for each individual and compared across different conditions (**Fig. 2A**). This analysis demonstrated a significant increase in the number of autoantibodies in patients with COVID-19, AD, AAV, SLE, and SSc compared to HCs.

**Figure 2.**
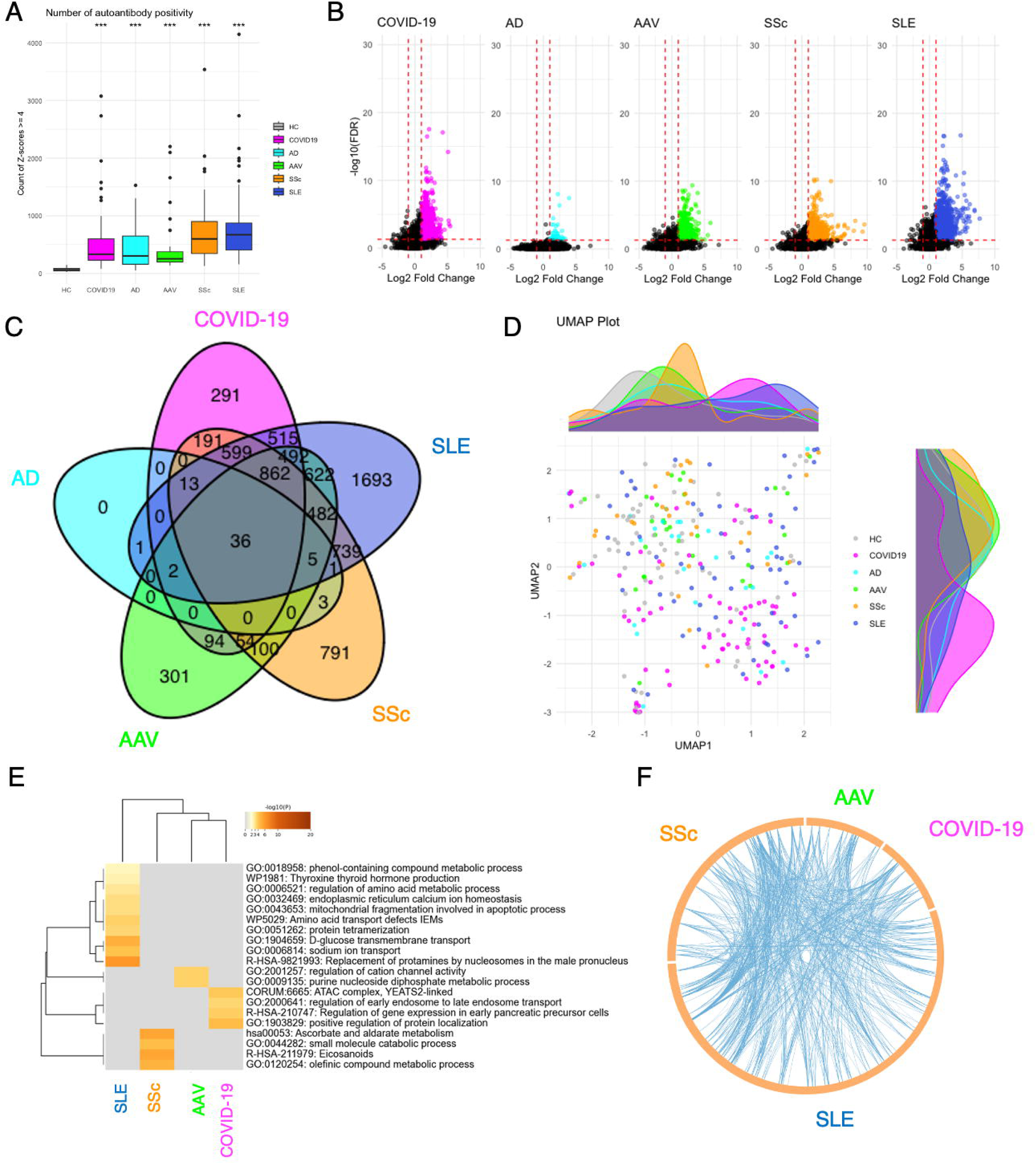
Identification of disease-specific autoantibodies. **(A)** Box plots that show the count of autoantibody positivity in each individual by different condition. ***: P < 0.001. P-values were calculated by two-sided Mann Whitney U test compared to HCs. **(B)** Volcano plots that illustrate differentially elevated autoantibodies within each condition compared to HCs. Red horizontal dash lines indicate false discovery rate (FDR) = 0.05. FDR was calculated by two-sided Mann Whitney U test with Benjamini-Hochberg correction compared to HCs. Red vertical dash lines indicate Log2 Fold Change (Disease/HC) = ±1. **(C)** Venn diagram that demonstrates the subsumptions among disease-specific autoantibodies. **(D)** UMAP plot that illustrates the distribution of disease-specific autoantibody profiles of each individual. **(E)** Heat map that shows the result of enrichment analysis targeting the genes responsible for the proteins targeted by such disease-specific autoantibodies. P values were calculated utilizing the list of 13,550 genes included in our cDNA library used for WPA manipulation as a reference. **(F)** Circos plot that depicts the overlap of the gene lists responsible for the proteins targeted by the disease-specific autoantibodies at the biological function level.

### Identification of disease-specific autoantibodies

We identified distinct sets of autoantibodies that showed a more than twofold significant increase in each disease condition relative to HCs (**Fig. 2B**). Notably, certain autoantibodies were unique to each disease (**Fig. 2C**). To illustrate the variability in serum levels of these disease-specific autoantibodies across individuals, we utilized uniform manifold approximation and projection (UMAP). UMAP analysis demonstrated inter- and intra-disease heterogeneity in autoantibody profiles. However, the UMAP plots did not show clear clustering based on disease state, indicating substantial overlap among the different conditions (**Fig. 2D**). Gene ontology analysis linked to the genes responsible for the proteins targeted by such disease-specific autoantibodies pointed to shared biological functions, with a focus on viral infection pathways and cytokine signaling in immune system, in COVID-19, SLE, and AAV (**Fig. 2E and 2F**). Holistic analysis of autoantibodies targeting cytokines, or their receptors displayed on our WPAs revealed that strong positivity for autoantibodies targeting type 1 interferon was specifically observed in COVID-19 patients, while weak positivity was seen in SLE patients (**Extended Fig. 2**).

### Selection of machine learning frameworks

To further investigate the association between autoantibody profiles and COVID-19, we adopted a machine learning approach. We tested nine different methods to differentiate COVID-19 cases from the others: simple linear regression, Ridge regression, logistic regression with data normalization, logistic regression with data standardization, support vector machine (SVM) with data normalization, SVM with data standardization, and extremely gradient boosting decision trees (XGBoost). As a result, XGBoost showed the highest value of the area under the receiver-operator characteristics curve (ROC-AUC) and Matthew’s Correlation Coefficient (MCC) for distinction of COVID-19 cases from the others (**Extended Table 2**). Consequently, we opted to focus on this method for our subsequent analysis.

### Performance of XGBoost

In our subsequent analysis using the entire dataset, we experimented with binary (COVID-19 vs. others), ternary (mild COVID-19 vs. moderate to severe COVID-19 vs. others), and multiclass (mild COVID-19 vs. moderate to severe COVID-19 vs. AAV vs. AD vs. SSc vs. SLE vs. HCs) classifications through XGBoost. The models achieved high accuracy in both binary and ternary classifications and showed significantly better outcomes than chance in the complex seven-class classification (**Extended Table 3**). The most significant autoantibodies identified across all models are depicted in **Fig. 3A, 3B, and 3C**, with autoantibodies against translational products from *BCL6 Corepressor Pseudogene 1* (*BCORP1*) emerging as a top feature in every model. Similarly, antibodies against K-Acetyltransferase 2A (KAT2A) were consistently prominent. Notably, Anti-BCORP1 and anti-KAT2A Abs were highlighted as important items in all the candidate machine learning methods tested (**Extended Fig. 3**). There was a correlation between anti-BCORP Abs and anti-KAT2A Abs as illustrated in **Fig. 3D, 3E, and 3F**. Remarkably, established serum markers for SSc and SLE, such as anti-topoisomerase 1 (TOP1) Abs, anti-centromere protein-B (CENPB) Abs, anti-tripartite motif-containing protein 21 (TRIM21) Abs, anti-small nuclear ribonucleoprotein polypeptide (SNRP)-A Abs, and anti-SNRPB Abs, were also identified. The visualization of mean serum levels of these prominent markers through spider charts revealed distinctive patterns across the different conditions (**Fig. 3G, 3H, and 3I**). We also attempted to integrate clinical features, specifically sex and age, into the classifiers. As a result, while age ranked second in the 3-class classification task and in the fourth in the 7-class classification task (**Extended Fig. 4A, 4B, and 4C**), it did not lead to improvement in overall performance across all classification tasks (**Extended Table 3**).

**Figure 3.**
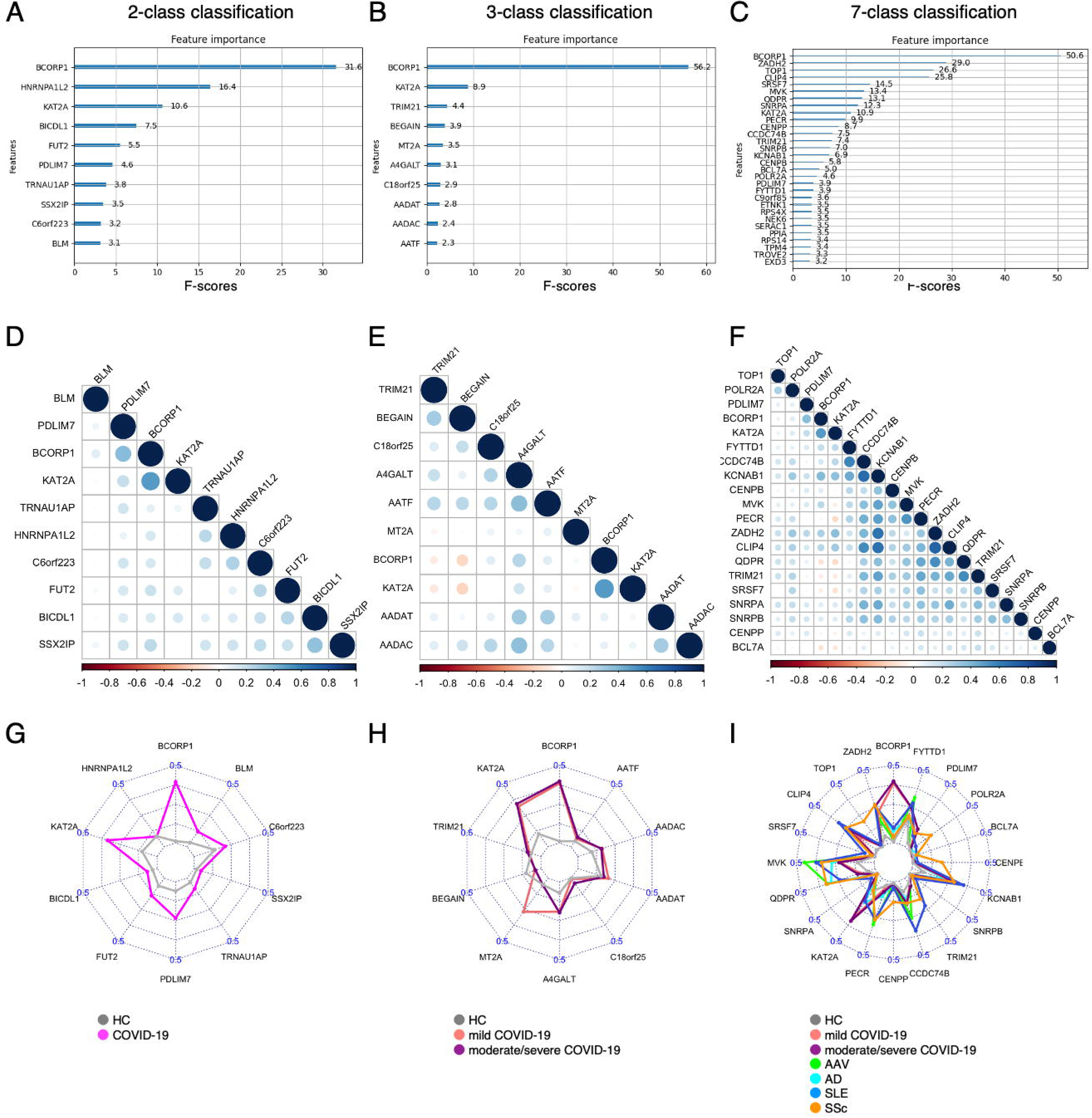
Autoantibodies highlighted in each machine learning model. Autoantibodies that are mostly highlighted according to feature importance in two-class **(A)**, three-class **(B)**, and multi-class **(C)** classifications. The feature importance is represented by F-scores, meaning the average number of splits by each feature over the cross-validation. Correlograms depict correlations, showcasing the connection between the highest-ranked autoantibodies in two-class **(D)**, three-class **(E)**, and multi-class **(F)** classifications. The correlation strength is denoted by Spearman’s rho on the color scale. Circle sizes represent the significance of the p-values, with only those with P < 0.05 being displayed. Radar charts present the average normalized quantities of the most important autoantibodies in each model, with line colors distinguishing between different disease categories in two-class **(G)**, three-class **(H)**, and multi-class **(I)** classifications.

### Minimum feature model

We evaluated the effectiveness of key features identified by machine learning algorithms for COVID-19 classification by training an XGBoost model during each iteration of cross-validation, using the top 1 to 5 features highlighted in the previous 2-class classification task (**Extended Fig.5**). Our analysis showed that even a single feature, BCORP1, achieves ROC-AUC exceeding 0.85, demonstrating strong discriminative power. While incorporating additional features enhanced some performance metrics, we observed a plateau beyond three features, suggesting that minimal models can still achieve high clinical relevance.

To further validate these findings, we trained a logistic regression model using the top two and top five features (**Extended Table 4**). Two-feature model (BCORP1 and KAT2A) achieved an AUC of 0.912, confirming that these markers are highly informative for COVID-19 classification. Five-feature model (BCORP1, KAT2A, RPS4X, BEGAIN, and TRIM21) improved accuracy, recall, precision, and F1-score. However, the ROC-AUC remained like that of the two-feature model. These results indicate that BCORP1 and KAT2A are key features for distinguishing COVID-19 cases, and our machine-learning approach is effective in identifying important biomarkers.

### Clinical relevance of autoantibodies

The serum levels of the top 20 autoantibodies highlighted through multi-class classification through XGBoost for each participant were depicted in a heatmap (**Fig. 4A**). Hierarchical clustering identified three unique groups of autoantibodies: cluster I, which included two autoantibodies highly specific to COVID-19 (anti-BCORP1 and anti-KAT2A Abs); cluster II, comprising autoantibodies that are commonly elevated across various conditions; and cluster III, involving well-established biomarkers for SSc and SLE. Principal component analysis (PCA) effectively distinguished between seven categories (**Fig. 4B**), particularly using principal component (PC) 2 as an indicator for COVID-19 (**Fig. 4C**). Correspondingly, antibodies against BCORP1 and KAT2A constituted the predominant contribution to PC2 (**Fig. 4D**).

**Figure 4.**
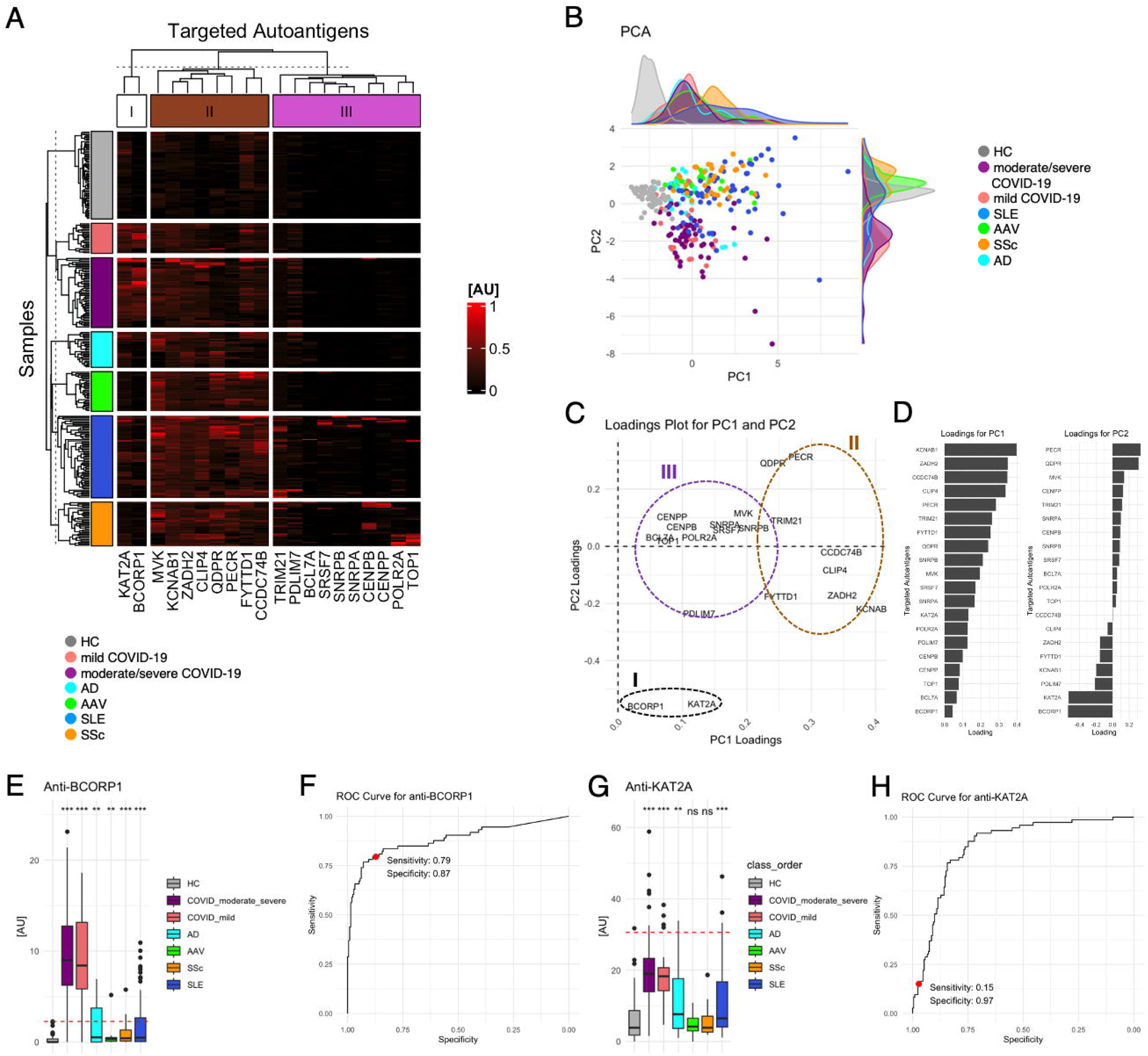
COVID-19 signature of autoantibody landscape. **(A)** The heatmap’s columns display the serum autoantibody concentrations highlighted in the multi-class classification using XGBoost. **(B)** PCA graph plots individual participants as points, with color coding to differentiate among various disease classes. **(C)** The loading diagram illustrates the contributions to PC1 and PC2, with I, II, and III marking the clusters defined in (A). **(D)** The bar graphs show the loadings of each autoantibody on PC1 and PC2. **(E)** A box plot presents the serum levels of anti-BCORP1 Abs in the subjects. **: P < 0.01, ***: P < 0.001. P-values were calculated by two-sided Mann Whitney U test compared to HCs. The red horizontal dash line indicates mean + 4SD in HCs. **(F)** A ROC curve illustrates the sensitivity and specificity for COVID-19 identification by serum levels of anti-BCORP1 Abs. The red dot marks the cutoff value set at mean + 4SD in HCs. **(F)** A box plot indicates the serum levels of anti-KAT2A Abs in the subjects. ns: P > 0.05, **: P < 0.01, ***: P < 0.001. P-values were calculated by two-sided Mann Whitney U test compared to HCs. The red horizontal dash line indicates mean + 4SD in HCs. **(G)** A ROC curve illustrates the sensitivity and specificity for COVID-19 identification by serum levels of anti-KAT2A Abs. The red dot marks the cutoff value set at mean + 4SD in HCs.

To validate the PWAbS results using WPAs in COVID-19 patients, we performed enzyme-linked immunosorbent assays (ELISA) to quantify serum levels of anti-KAT2A antibodies, employing recombinant KAT2A produced in a Baculovirus expression system. The analysis revealed a statistically significant positive correlation between the results obtained from PWAbS and ELISA (**Extended Fig. 6A**). In contrast, ELISA using recombinant BCL2 co-repressor (BCOR), which shares approximately 99% nucleotide sequence similarity with BCORP1 and was synthesized in HEK293 cells, showed no correlation with the serum levels of anti-BCORP1 antibodies measured by WPAs (**Extended Fig. 6B**).

Serum levels of anti-BCORP1 were the highest in COVID-19 among the examined conditions (**Fig. 4E**). This trend was consistent among both sex and age groups (**Extended Fig. 6C**). When the cutoff value was set at the mean + 4SD in healthy controls (HCs), only one individual in the HC group showed serum positivity for BCORP1, compared to 61 patients with COVID-19 (P < 0.001, two-sided Fisher’s exact test). Receiver operating characteristic (ROC) curve analysis demonstrated a sensitivity of 79% and a specificity of 87% (**Fig. 4F**). For anti-KAT2A antibodies, serum levels were highest in COVID-19 patients across all age and sex groups (**Figure 4G and Extended Fig. 6D**). With the cutoff set at the mean + 4SD in HCs, only one HC individual showed serum positivity for KAT2A, while 16 COVID-19 patients tested positive (P = 0.0045, two-sided Fisher’s exact test). ROC curve analysis indicated a sensitivity of 15% and a specificity of 97% (**Fig. 4H**). We also explored the link between COVID-19 clinical outcomes and the presence of anti-BCORP1 or anti-KAT2A Abs, but no significant association was found (**Extended Table 5 and 6**).

### Time course of autoantibody levels during COVID-19

Finally, we enhanced our analysis by including “late” time point samples. The data presented thus far derive exclusively from “early” time points, defined as within 10 days of symptom onset. To conduct a longitudinal evaluation of the humoral immune response, we incorporated paired serum samples from 41 individuals, including both “early” time points and “late” time points, defined as 11–20 days after symptom onset. These samples were used for PWAbS and for assessing IgG levels against SARS-CoV-2 particles: the nucleocapsid protein (N), spike protein (S), and the receptor binding domain (RBD) of S. Consistent with our prior findings, early timepoint samples showed IgG against N, S, and RBD in only a small part of the patients, with a marked increase in most patients by the late timepoint (**Fig. 5A**). To the contrary, SAL remained unchanged over time (**Fig. 5B**). Further exploration revealed a significant decrease in 293 autoantibodies and increase in 116 autoantibodies over the course of the infection, including those targeting BCORP1 and KAT2A (**Fig. 5C and 5D**). There was no observed correlation between these autoantibodies and IgG levels against N, S, and RBD (**Fig. 5E**). Notably, both anti-BCORP1 and anti-KAT2A Ab levels rose over time, especially in mild COVID-19 group for anti-BCORP1 and in moderate to severe COVID-19 group for anti-KAT2A (**Fig. 5F**). Additional analysis did not find any correlation between these two autoantibodies and IgG targeting N, S, and RBD at either timepoint or their progression over time (**Fig. 5G and 5H**).

**Figure 5.**
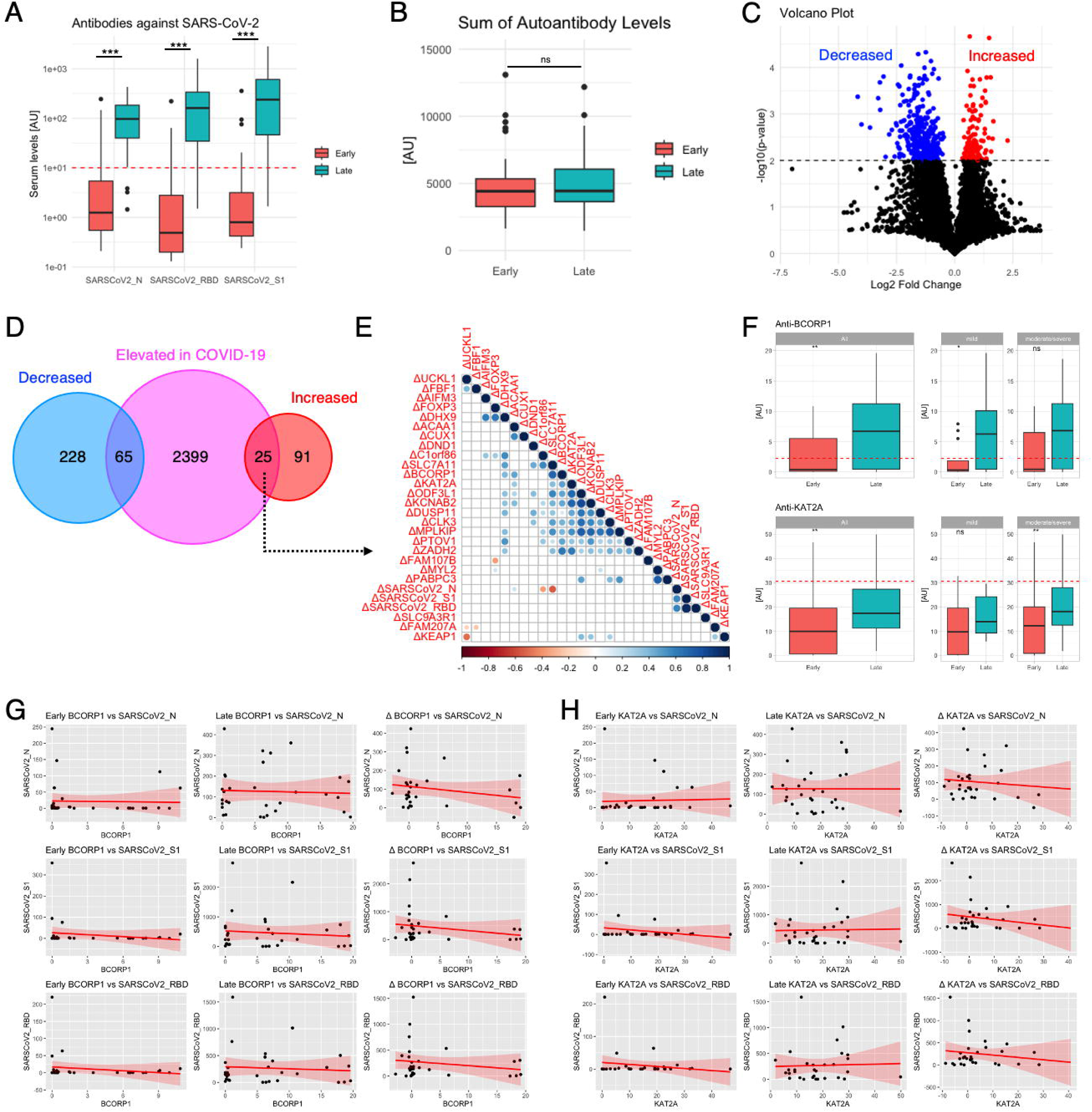
Longitudinal change of humoral immune response in COVID-19. **(A)** This box plot outlines the serum concentrations of IgG antibodies against N, S, or RBD in patients with COVID-19 at two intervals: “early” signifies within 10 days of symptom onset, and “late” refers to 11-20 days after symptoms appear. A red dashed line marks the threshold for a positive test result. ***: P < 0.001. P-values were calculated by two-sided Wilcoxon singed rank test. **(B)** These box plots demonstrate the SAL in patients with COVID-19 during the “early” and “late” time points. ns: P > 0.05. P-values were calculated by two-sided Wilcoxon singed rank test. **(C)** The volcano plot highlights biomarkers that are significantly increased (in red) or decreased (in blue) over time in patients with COVID-19. **(D)** A Venn diagram illustrates the overlap between autoantibodies that changed over time and those specific to COVID-19. **(E)** The correlogram visualizes the relationships between the overlapping autoantibodies that either increased over time or are specific to COVID-19, with the color scale indicating the strength of correlation according to Spearman’s rho and circle sizes depicting the significance of p-values, focusing on those with P < 0.05. **(F)** These box plots depict the time-based evolution of serum levels of anti-BCORP1 and anti-KAT2A antibodies in COVID-19 patients, categorized by disease severity. ns: P > 0.05, *: P < 0.05, **: P < 0.01. P-values were calculated by two-sided Wilcoxon singed rank test. **(G)(H)** The scatter plots illustrate the correlation between serum levels of anti-BCORP or anti-KAT2A Abs and IgG antibodies against the COVID-19 components at different time points, as well as their progression over time. The red lines and the surrounding shaded areas indicate the regression line and the 95% confidence interval, respectively.

## Discussion

Herein we conducted PWAbS subjecting serum samples derived from HCs and patients with COVID-19, AD, AAV, SLE, and SSc (**Fig. 1A**). We demonstrated that our PWAbS methodology enables us to identify disease-specific autoantibodies (**Fig. 2A, 2B, and 2C**), which demonstrate the distinct distribution of autoantibodies and common biological processes among different conditions (**Fig. 2D, 2E, and 2F**). The contrast in autoantibody profiles was accentuated through the application of a machine learning approach, particularly leveraging the XGBoost framework (**Fig. 3 and 4**). We also investigated the longitudinal change of autoantibody profiles along with the time course of COVID-19 and its correlation with emergence of antibodies targeting COVID-19 particles (**Fig. 5**). Collectively, these results supported our hypothesis that the combination of PWAbS and omics-based bioinformatic methodologies is adaptable to human disorders including COVID-19. Additionally, our findings in this study as well as our previous works provide a comprehensive catalog of autoantibody profiles in various diseases and open the door to creating innovative diagnostic methods that can differentiate between various disease mechanisms affecting multiple organs, utilizing distinct autoantibody patterns measured by WPAs.^17,19^

The machine learning-based approach has discovered that the presence of anti-KAT2A antibodies is highly specific to COVID-19 (**Fig. 4G and 4H**). The levels of anti-KAT2A Abs increased over the course of COVID-19 but were not correlated to antibodies targeting SARS-CoV-2 particles (**Fig. 5H**). This observation indicates that autoantibodies to KAT2A emerge because of autoantigen exposure due to tissue damage triggered by COVID-19, not as a reflection of cross-reaction between SARS-CoV-2 virions. KAT2A functions as a histone acetyltransferase that plays a role in the epigenetic regulation of the genome by modifying chromatin structures. There is notable research by John K. et al., indicating that the SARS-CoV-2 ORF8 protein mimics the histone H3’s ARKS motifs, which interfere with the role of KAT2A role in host cell epigenetic regulation.^28^ Intriguingly, the results of ELISA utilizing recombinant KAT2A produced in a Baculovirus expression system showed statistifcally significant positive correlation with our PWAbS results (**Extended Fig. 6A**), supporting the validity of our method for quantifying autoantibodies across different protein synthesis systems.

Anti-BCORP1 antibodies were also found to be specifically elevated in the sera of COVID-19 patients (**Fig. 4E and 4F**). Despite *BCORP1* being categorized as a pseudogene, it appears to undergo transcription into mRNA, as several transcriptomic studies have reported the presence of *BCORP1*-derived sequences.^29,30^ Especially, Deng MC’s research has shown that the transcription levels of *BCORP1* in peripheral blood mononuclear cells of COVID-19 patients correlate with early functional recovery and 1-year survival.^30^ However, it remains unclear whether *BCORP1* mRNA is translated into functional proteins, and further investigation is needed in this regard. Moreover, our discovery of anti-BCORP1 Abs in both males and females raises questions, as BCORP1 is located on the Y chromosome. This leads us to consider the possibility of cross-reactivity of these antibodies against foreign antigens, such as proteins comprising SARS-CoV-2 virions. However, we could not find any correlation between serum levels of anti-BCORP1 Abs and antibodies targeting SARS-CoV-2 particles at any of the timepoints examined, nor in their changes over time (**Fig. 5G**). Another hypothesis is cross-reaction between the antigen we produced from *BCORP1* cDNA and other human proteins. The *BCOR* gene, the counterpart of *BCORP1* found on the X chromosome, has a nucleotide sequence that is over 99% identical to *BCORP1*. As our cDNA library lacked BCOR cDNA, we used recombinant BCOR synthesized in HEK293 cells to perform ELISA for anti-BCOR antibody detection. The results showed no correlation with anti-BCORP1 antibody measurements obtained via WPA (**Extended Fig. 6B**), suggesting two possibilities: (1) the autoantibodies in COVID-19 target a different region of BCORP1 than BCOR, or (2) the autoantibodies recognize a shared region, but differences in their three-dimensional structures—potentially due to the recombinant BCOR not being preserved in a hydrated state—resulted in dissociation.

Our study has multiple strengths. First, the wheat-germ *in vitro* protein synthesis system and technique for manipulation of WPAs realized high-throughput expression of various human proteins including exoproteome upon a single platform. Second, as a result, our autoantibody measurement could cover a wider range of antigens at an almost proteome-wide level, which enabled us to apply omics-based bioinformatical approaches for interpreting the data. Third, engagement of machine learning methodologies enabled us to identify autoantibodies highly specific to COVID-19. Fourth, we investigated the longitudinal change of autoantibody profiles within COVID-19 patients, along with the measurement of antibodies targeting SARS-CoV-2 particles.

The limitation of our present study includes its retrospective design and a relatively small number of the subjects. The demographic differences, such as sex and age, between COVID-19 patients and other groups, including HCs, raise concerns about potential confounding, as previous studies have linked age and sex to autoantibody profiles.^31–33^ Additionally, it is important to note that critically ill patients were included only in the COVID-19 group, and elevated autoantibodies have been reported in critically ill individuals without COVID-19.^34^ Furthermore, we could not distinguish whether the autoantibodies found in our measurement were predisposed before COVID-19 or newly appeared after infection. In addition, functional assays for the autoantibodies such as neutralizing assays or *in vivo* studies are lacking. Therefore, insights into the direct contribution of each autoantibody to the pathophysiology of COVID-19 are limited. Our next challenges would include collecting serum samples before and after COVID-19 by accessing to population-based cohorts, recruiting sex, age, and severity-matched controls, evaluating the function of each autoantibody against their target molecules, and testing their contribution to the pathogenesis in animal experiments. From a machine learning aspect, there is another limitation caused by the small amount of data and imbalanced data. The small and imbalanced data may lead our model to fail to capture important features to discriminate against COVID-19 patients. This concern often arises in material science and life science. Nevertheless, in material science, it is revealed that various machine learning techniques, including XGBoost, are effective for such data^35^. Similarly, in life science, Parkinson’s disease specific features are identified from small and imbalanced data^36^. Therefore, in this study, various machine learning techniques we used are also considered to be effective in identifying the specific features of COVID-19 patients. To be more persuasive, we investigate the effectiveness of our discovered features from a clinical aspect instead of increasing the samples. As our future work, it is important to verify the discovered features’ effectiveness using more samples.

In conclusion, our study highlights the utility of PWAbS combined with bioinformatics-based autoantigenomics in identifying disease-specific autoantibody signatures. The findings not only provide insights into the unique autoantibody profiles of COVID-19 but also underscore the potential of this approach in understanding immune dysregulation across various diseases. By leveraging machine learning, we identified anti-BCORP1 and anti-KAT2A antibodies as highly specific markers for COVID-19, paving the way for innovative diagnostic methods. Validation of these findings in external cohorts will be crucial to confirm their generalizability and clinical utility, as well as to further explore the functional roles of these autoantibodies in disease mechanisms.

## Acknowledgements

We thank Ms. Maiko Enomoto and her colleagues for secretary work. We thank Ms. Teruko Tani and Ms. Mayumi Odagiri for their assistance in clinical data collection. We appreciate Ms. Maiko Matsuda, VESPER Studio Inc., Tokyo, Japan, for her contribution of illustrating skills.

## Author Contributions

KM Matsuda primarily engaged in autoantibody measurement, clinical data collection, data analysis, visualization, and writing the first draft of the manuscript. Y Kawase primarily contributed to machine learning analysis. K Iwadoh also participated in machine learning analysis. M Kurano, Y Yatomi, K Okamoto, and K Moriya participated in sample collection and clinical data acquisition regarding COVID-19. H Kotani oversaw ELISA experiments. A Kuzumi, T Fukasawa, A Yoshizaki-Ogawa took part in the sample collection of SSc. T Hisamoto was in charge of sample collection of AD. M Kono, T Okamura, H Shoda, and K Fujio oversaw sample collection of SLE. K Yamaguchi, T Okumura, C Ono, Y Kobayashi, A Sato, A Miya, and N Goshima prepared wet protein arrays, provided technical assistance for autoantibody measurement, participated in data analysis, setup of UT-ABCD, and revised the manuscript. R Uchino, Y Murakami and H Matsunaka provided technical assistance for autoantibody measurement. H Imai and R Raymond supervised the study. S Sato conceptualized and supervised the study. A Yoshizaki conceptualized, launched, and supervised this study, and was involved in revising the manuscript.

## Data availability statement

The digest of the results is available as “aUToAntiBody Comprehensive Database (UT-ABCD)” at http://www.ut-abcd.org. The full dataset is available upon reasonable request to the corresponding author, in compliance with ethical guidelines and participant privacy considerations.

## Materials and Methods

### Human subjects

We consecutively enrolled patients administered to our institution for COVID-19 from April 2020 to April 2021. Inclusion criteria were a SARS-CoV-2 positive nasopharyngeal swab test by real-time reverse transcription-polymerase chain reaction (RT-PCR) and age ≥18 years. A Ct value of 38 cycles was used as the threshold for determining a positive result in RT-PCR. Clinical data were collected by retrospective review of electric medical records. We gathered basic patient information, symptoms, medications, histopathologic features, and laboratory findings from the closest time point from the date of serum collection. The disease severity was assessed following the Japanese guideline for managing COVID-19 patients.^37^ In brief, individuals requiring intensive care or mechanical ventilation were categorized as severe COVID-19, those exhibiting hypoxemia among the remaining cases were classified as moderate to severe COVID-19, and all other patients were considered to have mild COVID-19. Data were derived from samples collected at an “early” time point, defined as within 10 days of symptom onset. Additionally, for Figure 5, we included samples from the same COVID-19 patients collected at a “late” time point (11–20 days after symptom onset) in 41 patients, where available. We also gathered serum samples from HCs and patients with AD, AAV, SLE, and SSc. HCs were randomly recruited from healthcare providers with no medical history, who underwent annual health checkups. Patients with AD, AAV, SLE, and SSc were recruited from individuals regularly visiting The University of Tokyo Hospital, all of whom met the diagnostic or classification criteria for their respective conditions.^38–41^ This study has been approved by The University of Tokyo Ethical Committee (Approval number 0695). Written informed consent has been obtained from all the participants.

### Measurement of IgG targeting SARS-CoV-2 particles

The process of quantifying IgG antibodies that target specific SARS-CoV-2 proteins, namely the nucleocapsid protein, spike protein, and the spike protein’s receptor binding domain, was conducted as outlined previously using a commercial SARS-CoV-2 IgG kit (YHLO Biotechnology Company, Ltd., Shenzhen, China).^42,43^ This involved an assay where serum samples were combined with magnetic beads coated with the viral proteins and a substance to prepare the samples. This mix was then washed, combined with an acridinium-conjugated anti-human IgG, and washed again. The subsequent steps included adding solutions to induce a chemiluminescent reaction, the intensity of which was measured by the iFlash3000 CLIA analyzer (YHLO Biotechnology Company, Ltd.) A threshold of 10 AU/mL was used for the detection, following the guidelines provided by the manufacturer.

### Autoantibody measurement

WPAs were arranged as previously described.^17^ First, proteins were synthesized *in vitro* utilizing a wheat germ cell-free system from 13,352 clones of the HuPEX.^20^ POLR3A and TRIM21 existed in two forms: full-length and truncated. All other proteins were full-length. Second, synthesized proteins were plotted onto glass plates (Matsunami Glass, Osaka, Japan) in an array format by the affinity between the GST-tag added to the N-terminus of each protein and glutathione modified on the plates. All autoantigens were presented in duplicate on the WPAs. The array also included mock spots as negative controls (wheat germ cell-free protein synthesis products without any cDNA) and IgG spots as positive controls (wheat germ cell-free protein synthesis products with IgG-coding cDNA). The WPAs were treated with human serum diluted by 1:333 in the reaction buffer containing 1x Synthetic block (Invitrogen), phosphate-buffered saline (PBS), and 0.1% Tween 20. Next, the WPAs were washed, and goat anti-Human IgG (H+L) Alexa Flour 647 conjugate (Thermo Fisher Scientific, San Jose, CA, USA) diluted 1000-fold was added to the WPAs and reacted for 1 hour at room temperature. Finally, the WPAs were washed, air-dried, and fluorescent images were acquired using a Typhoon imaging system (Cytiva, Marlborough, MA, USA). Fluorescence images were analyzed to quantify serum levels of autoantibodies targeting each antigen, following the formula shown below:

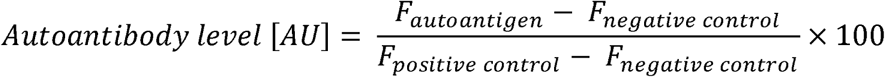

*AU*: arbitrary unit

*F _autoantigen_*: fluorescent intensity of autoantigen spot

*F _negative control_*: fluorescent intensity of negative control spot

*F _positive control_*: fluorescent intensity of positive control spot

When there was a significant discrepancy between the results of duplicate spots, we examined the raw fluorescence images and excluded abnormal signals, such as noise generated by dust particles. SAL was defined as the sum of serum levels for all 13,352 autoantibodies tested in our study.

### Machine learning

We trained supervised machine learning models for classification tasks using simple linear regression, Ridge regression, logistic regression with normalization, logistic regression with standardization, SVM with normalization, SVM with standardization, and XGBoost. These models were implemented using the scikit-learn library (https://scikit-learn.org/) in Python (v.3.11.11) and trained within a Jupyter Notebook environment. Hyperparameter tuning was conducted using Optuna (https://optuna.org/).

For simple linear regression, Ridge regression, and logistic regression, we used the “sklearn.linear_model” module. For Ridge regression, the hyperparameter “alpha” was searched between 0.0001 and 10.0 on a logarithmic scale, and the optimal value was determined to be 9.994. For logistic regression, we performed standardization or normalization of the data using the “sklearn.preprocessing” module.

For SVM, implemented using the “sklearn.svm” module, we applied standardization or normalization with the “sklearn_preprocessing” module, and feature importance was assessed by averaging the number of splits each feature contributed to over cross-validation.

For XGBoost, we used the “xgboost” module and performed hyperparameter tuning over the following ranges: “n_estimators” (100–1000), “max_depth” (3–9), “learning_rate” (1e-3–1e-1) on a logarithmic scale, “subsample” (0.6–1.0), “colsample_bytree” (0.6–1.0), “min_child_weight” (1–10), “reg_lambda” (1e-8–10.0) on a logarithmic scale, and “reg_alpha” (1e-8–10.0) on a logarithmic scale. The best hyperparameters obtained were “n_estimators”: 556, “max_depth”: 4, “learning_rate”: 0.003, “subsample”: 0.939, “colsample_bytree”: 0.635, “min_child_weight”: 8, “reg_lambda”: 0.370, and “reg_alpha”: 2.210.

### Performance Metrics

We performed the 10-fold cross-validation as follows: first, we shuffled and split the data into 10 equally sized parts, selecting one of these parts as validation data while the remaining 9 parts as training data. We then repeated this procedure 10 times, selecting a different part as the validation data. Model performance on the testing set was evaluated using the following metrics:

- Accuracy: 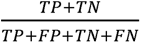
- Precision: 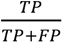
- Recall: 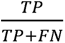
- F1-score: 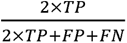
- Matthew’s Correlation Coefficient (MCC): 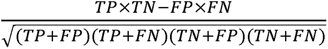

TP: true positive

FP: false positive

TN: true negative

FN: false negative

### Feature importance scores and feature selection

The feature importance was indicated by their mean absolute coefficient in the 10-fold cross validation in simple linear regression, Ridge regression, and logistic regression. As for SVM and XGBoost, the feature importance was represented by F-scores, meaning the average number of splits by each feature over the cross-validation. When constructing minimum feature models, we selected only the top 1–5 features ranked by F-scores for final model training in each cross-validation iteration. In all other cases, we analyzed either all measured autoantibodies alone or in combination with age and sex.

### Enzyme-Linked Immunosorbent Assay

Two 96-well antigen-binding plates were prepared: one for antigen coating and another for negative control. Each well was initially filled with 50 µL of phosphate-buffered saline (PBS). After incubation for 5 minutes at room temperature (RT), the liquid was removed. A solution of recombinant KAT2A (MyBioSource, San Diego, CA, USA) or recombinant BCOR (OriGene Technologies, Rockville, MD, USA) was prepared at a concentration of 3 µg/mL in PBS. Subsequently, 30 µL of the antigen solution was added to each well of the antigen coating plate, while 30 µL of PBS alone was added to the wells of the negative control plate. The plates were then incubated at 300 rpm for 1.5 hours at RT to facilitate antigen binding. After incubation, the plates were washed three times with 120 µL of PBST (PBS containing 0.1% Tween-20) per well. Blocking was performed by adding 100 µL of blocking buffer (0.5% skim milk in PBST) to each well. The plates were sealed with transparent adhesive film and incubated at 300 rpm for 1 hour at RT. Following blocking, adjusted serum reaction solutions were prepared and pipetted into the plates, with 50 µL added to each well. The plates were sealed with aluminum adhesive film and incubated at 860 rpm for 1 hour at RT. The plates were washed three times with 120 µL of TBST (Tris-buffered saline containing 0.1% Tween-20). Subsequently, 50 µL of secondary antibody solution, goat anti-Human IgG (H+L) Alexa Flour 647 conjugate (Thermo Fisher Scientific) diluted 1:1000 in TBST, was added to each well. The plates were again sealed and incubated for 1 hour at RT. After the secondary antibody reaction, the plates were washed three times with 120 µL of TBST. Then, 50 µL of TBST was added to each well. Fluorescence signals were scanned using a Typhoon imaging system (Cytiva). The serum autoantibody levels were quantified by calculating the difference in fluorescent signals between the antigen-coated plate and the negative control plate.

### Statistical analysis

Two-group comparisons were performed using two-sided Fisher’s exact test for categorical variables and two-sided Mann-Whitney U test for continuous variables. P-value of < 0.05 was considered statistically significant. Differentially elevated autoantibodies were defined as more than 2-fold changes in the serum levels with a false discovery rate (FDR) < 0.05, adjusted using the Benjamini-Hochberg method. Gene Ontology Analysis using web-based tools targeted the list of the entry clones coding the differentially highlighted autoantigens was performed for gene-list enrichment analysis, gene-disease association analysis, and transcriptional regulatory network analysis with Metascape.^44^ As a reference, we utilized the list of all the genes included in our cDNA library used for WPA manipulation. Other data analyses and presentations were conducted using Stata IC/15.0 (StataCorp, TX, USA).

### Data visualization

Box plots, scatter plots, hierarchical clustering and correlation matrix were visualized by using R (v4.2.1). Box plots were defined as follows: the middle line corresponds to the median; the lower and upper hinges correspond to the first and third quartiles; the upper whisker extends from the hinge to the largest value no further than 1.5 times the interquartile range (IQR) from the hinge; and the lower whisker extends from the hinge to the smallest value at most 1.5 times the IQR of the hinge.

## Code availability

All the scripts are available at https://github.com/mkazukikom/COV19ML.

**Extended Figure 1. The sum of autoantibody levels by sex and age. (A)** A box plot illustrates sum of autoantibody levels (SAL) in each individual by different condition. SAL was defined as the sum of serum levels for all 13,352 autoantibodies tested in our study. ns: P > 0.05, ***: P < 0.001. P-values were calculated by two-sided Mann Whitney U test compared to HCs. **(B)** Box plots show SAL by sex and age groups. P-values were calculated by two-sided Mann Whitney U test compared to HCs. **(C)** A jitter plot displays the distribution of Z-scores for each measured autoantibody in healthy controls (HCs), arranged in descending order of their maximum Z-score values. The red horizontal dashed line marks Z-score = 4. **(D)** A histogram illustrates the distribution of Z-scores for all autoantibody measurements in HCs, with a red vertical dashed line marking Z-score = 4. **(E)** A cumulative histogram illustrates the distribution of Z-scores for all autoantibody measurements in HCs, with a red vertical dashed line marking Z-score = 4 and a red horizontal dashed line indicating 99%.

**Extended Figure 2. Autoantibodies to cytokines or their receptors. (A)** The heatmap’s columns display the serum autoantibody concentrations targeting cytokines in each subject evaluated by our proteome-wide autoantibody screening. **(B)** The heatmap’s columns display the serum autoantibody concentrations targeting cytokine receptors in each subject evaluated by our proteome-wide autoantibody screening. **(C)** A box plot presents the serum levels of anti-interferon alpha 2 (IFNA2) Abs in the subjects. **(D)** Another box plot indicates the serum levels of anti-interferon alpha 4 (IFN4A) Abs in the subjects.

**Extended Figure 3. Feature importance of top highlighted autoantibodies in other candidate machine learning frameworks for 2-class classification. (A)** Simple linear regression. **(B)** Ridge regression. **(C)** Logistic regression with normalization. **(D)** Logistic regression with standardization. **(E)** SVM with normalization, and **(F)** SVM with standardization. For simple linear regression, Ridge regression, and logistic regression, the feature importance was indicated by their mean absolute coefficient in the 10-fold cross validation. For SVM, the feature importance is represented by F-scores, meaning the average number of splits by each feature over the 10-fold cross-validation.

**Extended Figure 4. Integration of ciilical features.** Autoantibodies and clinical features that are mostly highlighted according to feature importance in two-class **(A)**, three-class **(B)**, and multi-class **(C)** classifications by XGBoost. The feature importance is represented by F-scores, meaning the average number of splits by each feature over the cross-validation.

**Extended Figure 5. Performance of minimal feature models based on XGBoost using the top 1–5 features.** The top-ranked 1–5 features identified by XGBoost, or all the measured autoantibodies, were utilized for a 2-class classification task to discriminate COVID-19 cases.

**Extended Figure 6. Serum levels of anti-BCORP1 and anti-KAT2A Abs by sex and age. (A)** A scatter plot depicts the correlation between serum levels of anti-KAT2A autoantibodies measured by wet protein arrays (WPAs) and enzyme-linked immunosorbent assays (ELISA). The red line represents the regression line, while the gray shaded area denotes the 95% confidence interval. AU: arbitrary unit. **(B)** A scatter plot depicts the correlation between serum levels of anti-BCORP1 autoantibodies measured by WPAs and serum levels of anti-BCOR autoantibodies by ELISA. The red line represents the regression line, while the gray shaded area denotes the 95% confidence interval. AU: arbitrary unit. **(C)** Serum levels of anti-BCORP1 Abs by sex and age groups. ns: P > 0.05, *: P < 0.05, **: P < 0.01, ***: P < 0.001. P-values were calculated by two-sided Mann Whitney U test compared to HCs. Red vertical dash lines indicate mean + 4SD in HC. **(D)** Serum levels of anti-KAT2A Abs by sex and age groups. ns: P > 0.05, *: P < 0.05, **: P < 0.01, ***: P < 0.001. P-values were calculated by two-sided Mann Whitney U test compared to HCs. Red vertical dash lines indicate mean + 4SD in HC.

