## Supplementary figures and images for "Proteome-wide autoantibody screening and holistic autoantigenomic analysis unveil COVID-19 signature of autoantibody landscape"

### Extended Figure 1

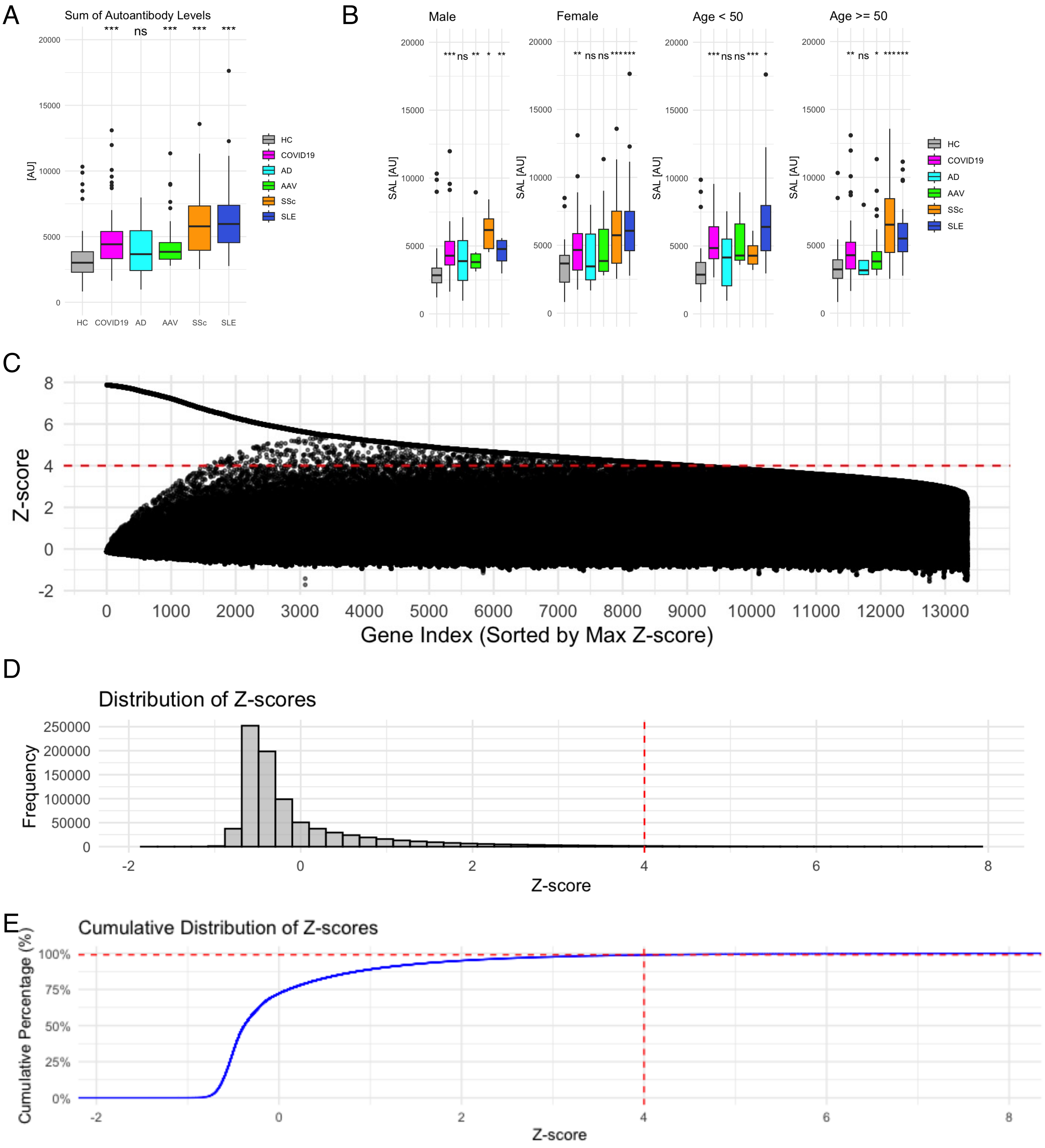


Matsuda KM et al.
Extended Figure 1

### Extended Figure 2

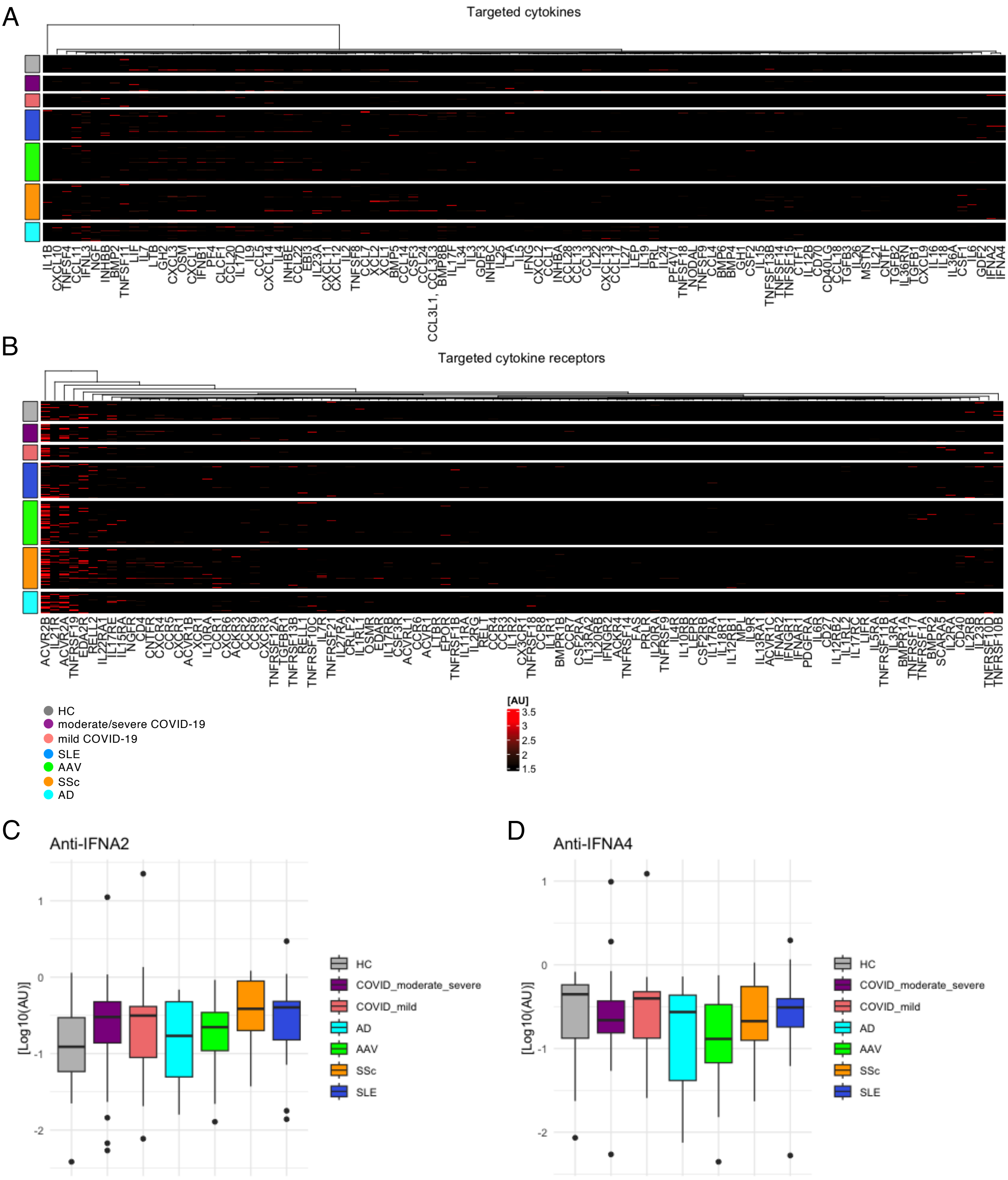


Matsuda KM et al.
Extended Figure 2

### Extended Figure 3

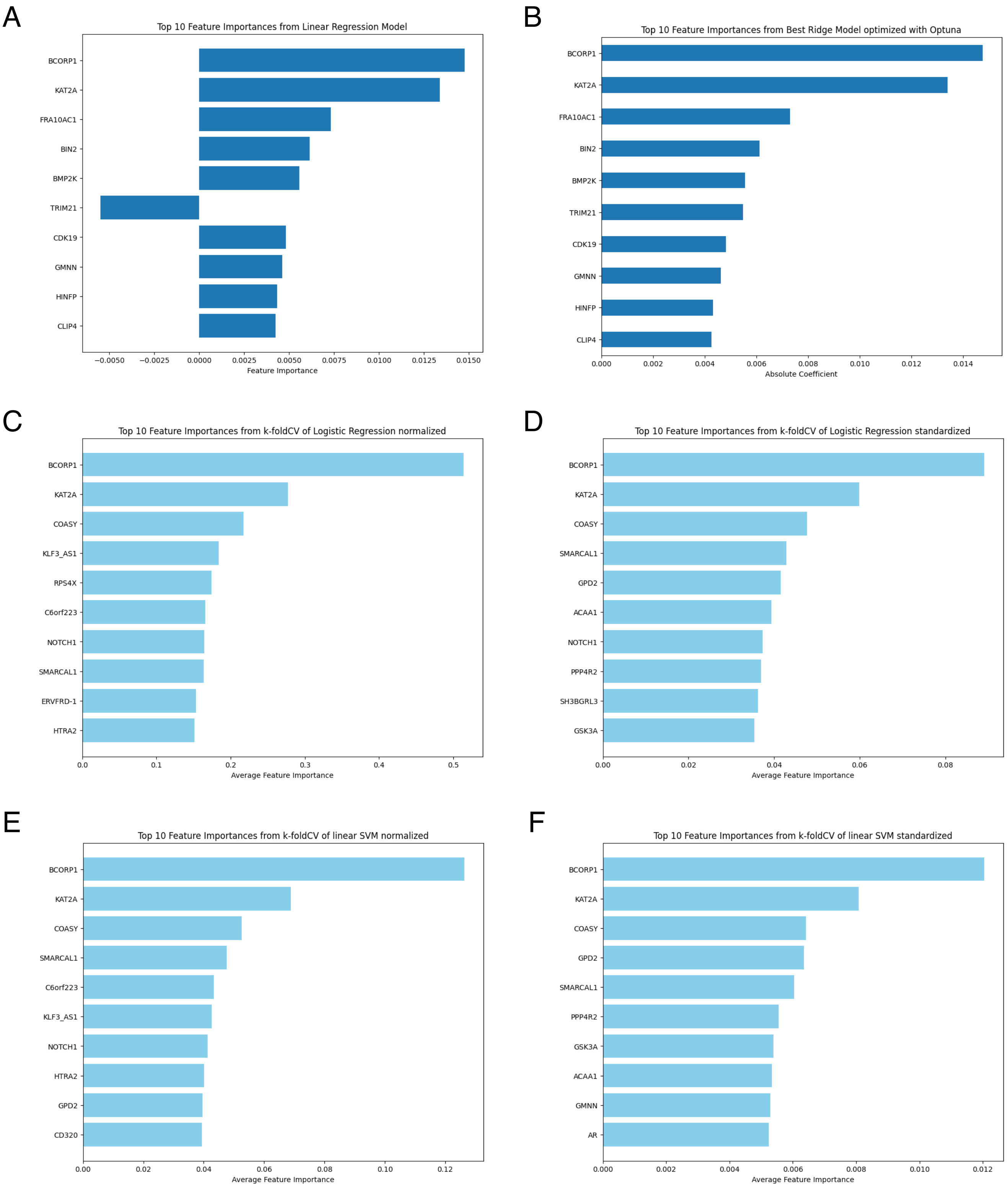


Matsuda KM et al.
Extended Figure 3

### Extended Figure 4

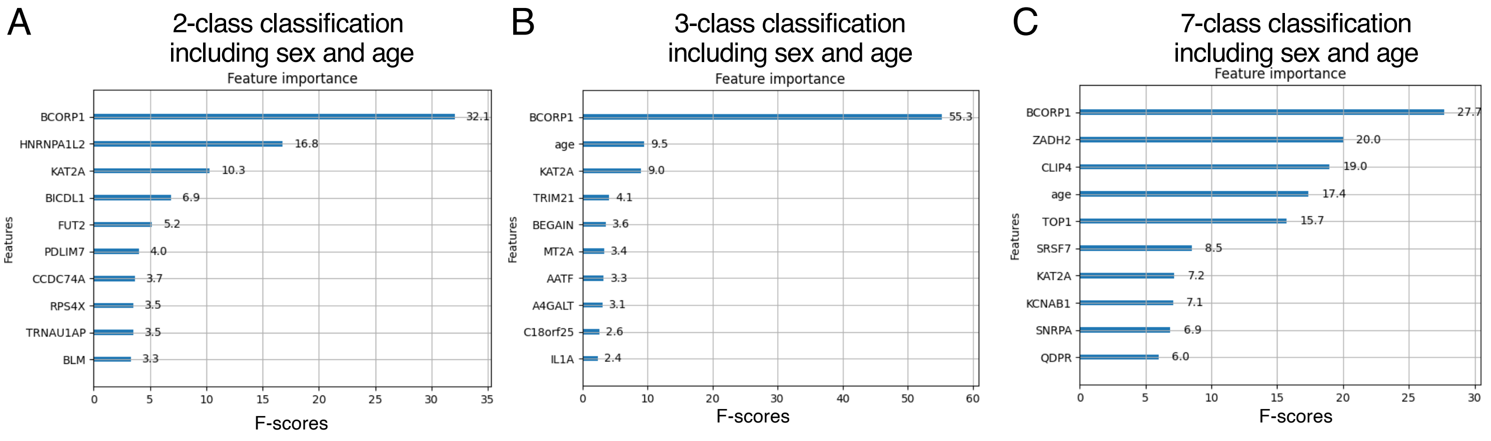


Matsuda KM et al.

Extended Figure 4

### Extended Figure 5

Performance of minimum feature models

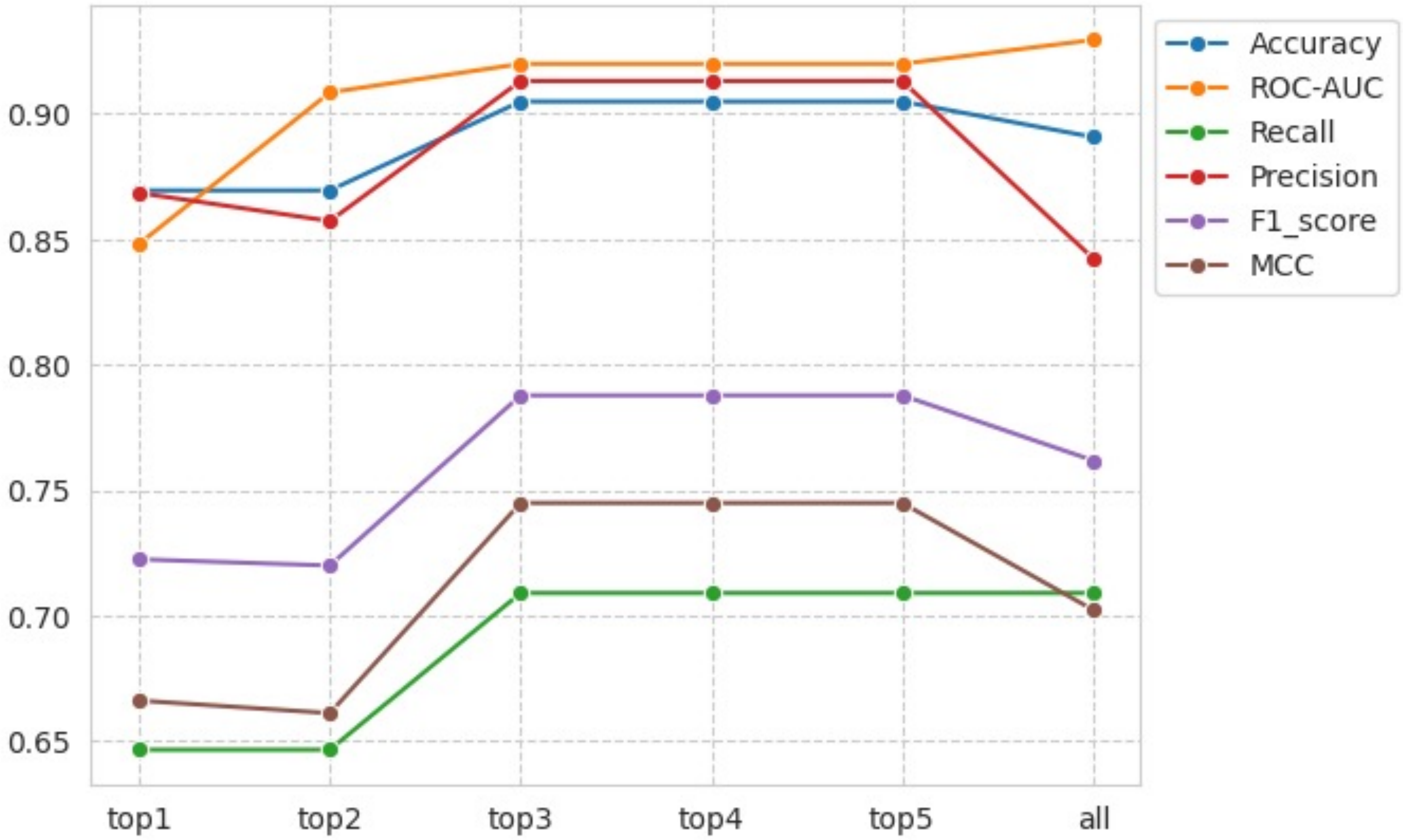


Matsuda KM et al.
Extended Figure 5

### Extended Figure 6

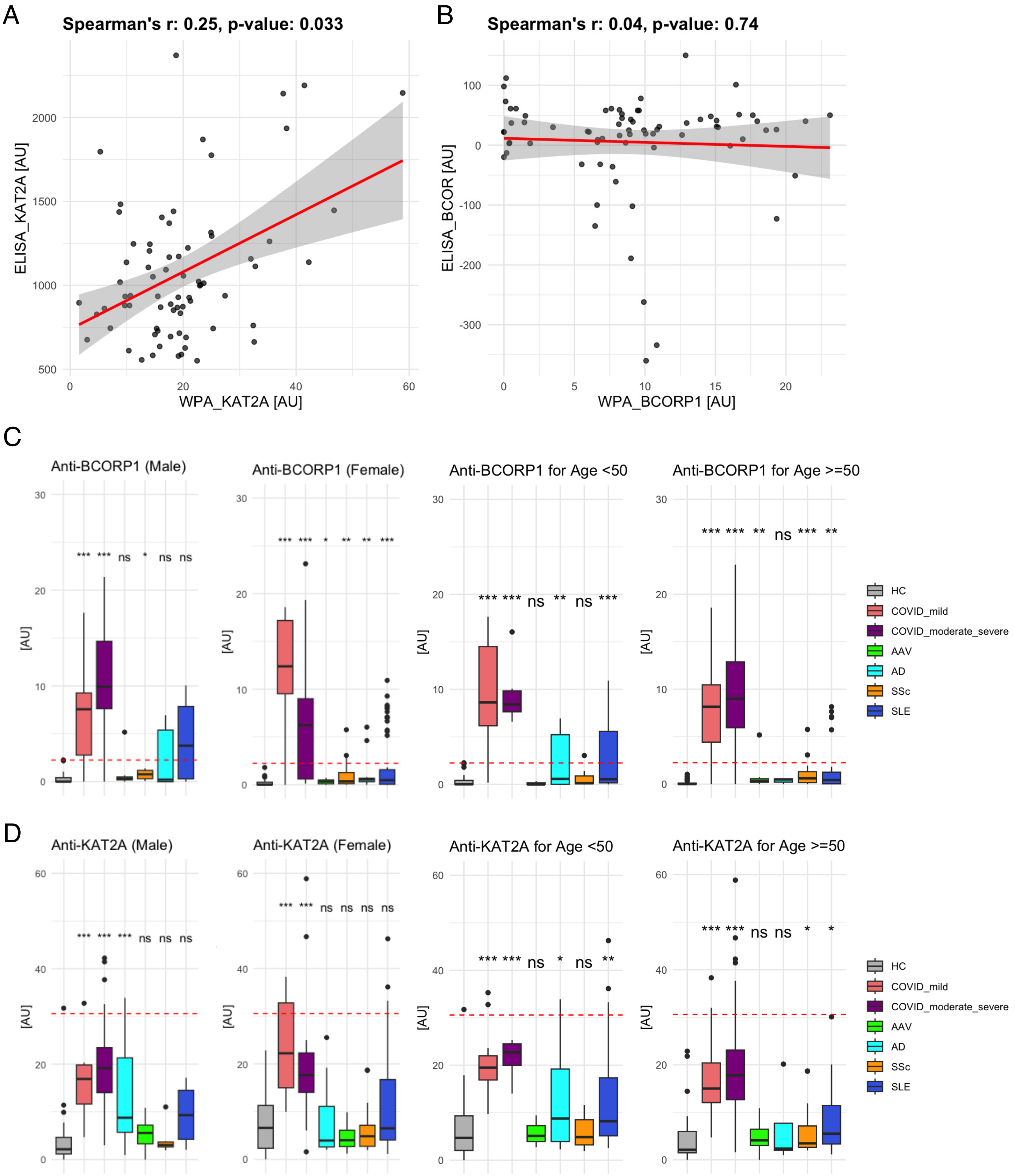


Matsuda KM et al.
Extended Figure 6
